# Prognostic value of visual quantification of lesion severity at initial chest CT in confirmed Covid-19 infection: a retrospective analysis on 216 patients

**DOI:** 10.1101/2020.05.28.20115584

**Authors:** Elias Taieb, Aïssam Labani, Yvon Ruch, François Danion, Pascal Bilbault, Mathieu Oberlin, Pierre Leyendecker, Catherine Roy, Mickaël Ohana

**Author notes:** **Corresponding author** Elias TAIEB, Nouvel Hôpital Civil - Service de Radiologie, 1 place de l’Hôpital, 67000 Strasbourg, FRANCE.

## Abstract

**Rationale and Objectives:** Studies suggest an association between chest CT findings assessed with semi-quantitative CT score and gravity of Covid-19. The objective of this work is to analyze potential correlation between visual quantification of lesion severity at initial chest CT and clinical outcome in confirmed Covid-19 patients.

**Materials and Methods:** From March 5^th^ to March 21^st^, 2020, all consecutive patients that underwent chest CT for clinical suspicion of Covid-19 at a single tertiary center were retrospectively evaluated for inclusion. Patients with lung parenchyma lesions compatible with Covid-19 and positive RT-PCR were included.

Global extensiveness of abnormal lung parenchyma was visually estimated and classified independently by 2 readers, following the European Society of Thoracic Imaging Guidelines. Death and/or mechanical ventilation within 30 days following the initial chest CT was chosen as the primary hard endpoint.

**Results:** 216 patients (124 men, 62 years-old ± 16.5, range 22 – 94 yo) corresponding to 216 chest CT were included. Correlation between lesion severity and percentage of patients that met the primary endpoint was high, with a coefficient ρ of 0.87 (p = 0.05).

A greater than 25% involvement was significantly associated with a higher risk of mechanical ventilation or death at 30 days, with a Risk Ratio of 5.00 (95% CI [3.59–6.99]).

**Conclusion:** This retrospective cohort confirms a correlation between visual evaluation of lesions severity at initial chest CT and the 30 days clinical outcome of Covid-19 patients and suggests using a threshold of greater than 25% involvement to identify patients at risk.

## INTRODUCTION

Covid-19 is due to the severe acute respiratory syndrome coronavirus 2 (SARS-CoV-2), first described in China in December 2019 and that has spread as a worldwide pandemic in March 2020. Even though most cases are benign, 17 to 29% of hospitalized patients may develop a severe pneumonia with acute respiratory distress syndrome (ARDS) necessitating aggressive resuscitative measures such as mechanical ventilation with prone position (1,2). Reverse-transcription polymerase chain reaction (RT-PCR) assay is the reference standard to confirm Covid-19 (3). Nonetheless, results are delayed and initial false negatives are frequent (4), resulting in the need for alternative triage methods in Emergency Department (ED) patients during high prevalence of the disease. Chest computed tomography (CT) is an established alternative for the early screening and triage of suspected Covid-19 patients with moderate to severe clinical symptoms. Indeed, chest-CT typical findings of Covid-19 pneumonia are now well described with ground-glass opacities (GGO), consolidation and crazy-paving pattern that are bilateral, multifocal with peripheral predominant distribution (4)(5).

Initial studies suggest an association between chest CT findings and gravity of Covid-19. Yuan et al. described on 27 consecutive patients a higher semi-quantitative CT score in the 10 that died compared to the 17 that recovered (6,7). Zhao et al. described on 101 cases a correlation between lesion severity on CT and clinical severity of the disease (8). Yang et al. also found on 102 Covid-19 patients a higher CT score in those with severe clinical disease (9). Correlation between initial chest-CT findings and a precise clinical outcome such as admission in intensive care unit (ICU) or mortality has been assessed in 236 patients by Colombi et al. They showed that software quantification of well-aerated lung parenchyma on initial chest-CT was predictive of ICU admission or death, with a suggested cut-off at 73% (10). This method would however require the use of dedicated quantification software, which is time-consuming and difficult to integrate in routine workflow. On the contrary, visual evaluation of lesion severity as recommended by the European Society of Thoracic Imaging Guidelines (11) is simple and easy to integrate in the everyday workflow. Yet, and to the best of our knowledge, no study has reported the potential predictive value of this approach.

The objective of this work is to analyze potential correlation between visual quantification of lesion severity at initial chest CT and the clinical outcome in confirmed Covid-19 patients.

## MATERIALS AND METHODS

This retrospective study was approved by the local ethics committee of Strasbourg University Hospital and the need of written informed consent was waived.

### Population

From March 5^th^ to March 21^st^, 2020, all consecutive patients that underwent chest CT for clinical suspicion of Covid-19 at our ED (Nouvel Hôpital Civil, Strasbourg University Hospital, France) were retrospectively evaluated for inclusion. Patients were addressed for moderate to severe respiratory symptoms and chest CT was systematically used at their ED arrival to expedite triage. Systematic RT-PCR sample obtained by nasopharyngeal swab were done concomitantly to the CT. Some patients could have a second or third sampling using expectorations or bronchoalveolar lavage if the first sample was negative.

Inclusion criteria were:

– availability of chest CT at the time of ED admission;
– presence of lung parenchyma lesions compatible with Covid-19 on the initial chest CT;
– at least one positive RT-PCR result.

Exclusion criteria were:

– delay between initial chest CT and the first positive RT-PCR greater than 14 days,
– lack of follow-up data at 30 days after initial chest CT.

### Clinical Evaluation

Death and/or mechanical ventilation within 30 days following the initial chest CT was chosen as the primary hard endpoint.

Age, sex, onset of symptoms at the time of chest CT and hospitalization status were retrieved from the Electronic Health Record.

### CT Acquisition Protocol

All CT examinations were acquired using an 80-row CT scanner (Aquilion PRIME SP, Canon Medical Systems), without injection of contrast media. Acquisition protocol was adapted according to the patient’s morphotype, with a tube voltage of 100–135kV and a tube current of 2–350mAs. Patients were instructed to hold their breath and raise their arms above their head to minimize artifacts. Images were reconstructed with a slice-thickness of 1 mm in mediastinal and parenchymal windows and transmitted to post-processing workstations for multiplanar and maximum intensity projection reconstructions.

### CT Image Analysis

Two radiologists (ET with a 3 years’ experience in CT and AL with 10 years’ experience) blinded to the clinical data, reviewed CT images independently and resolved discrepancies by consensus, after an initial training on a separate set of 20 cases.

Global extensiveness of abnormal lung parenchyma *- i.e*. lesions consistent with Covid-19 such as GGO, consolidation, crazy paving and organizing pneumonia – was visually estimated and classified following the European Society of Thoracic Imaging Guidelines as <10%, 11–25%, 26–50%, 51–75% or > 75% (11). This visual quantification was obtained after visual analysis of 5 zones on axial images: at the level of the supra-aortic trunks, the azygos cross, the carina, the inferior pulmonary veins and the pleural recesses.

### Statistical analysis

Continuous variables were expressed as median and standard deviation (SD). Normality of distribution was assessed using a Shapiro-Wilk test. Variables were compared using Student t-test when normally distributed, or Wilcoxon.

Categorical variables were expressed as counts and percentage, with corresponding 95% confidence interval (95% CI) using Wilson method, and compared using Pearson’s χ2 tests or Fisher’s exact tests.

Inter-reader agreement was assessed with a Cohen’s Kappa test.

Correlation between the severity group and the percentage of patients that met the primary endpoint was assessed with a Spearman test.

Risk ratio for primary endpoint was calculated between patients with less than 25% lesions and those with more than 25% lesions.

All the analyses were performed using R software version 3.6.3. (R Core Team (2019). R: A language and environment for statistical computing. R Foundation for Statistical Computing, Vienna, Austria. URL https://www.R-project.org/).

## RESULTS

### Population

598 patients were admitted in the ED for a clinical suspicion of Covid-19 over the inclusion period; 216 patients (124 men, 62 years-old ± 16.5, range 22 – 94yo) corresponding to 216 chest CT were finally retrospectively included in this work **(Fig 1)**.

**Fig 1.**
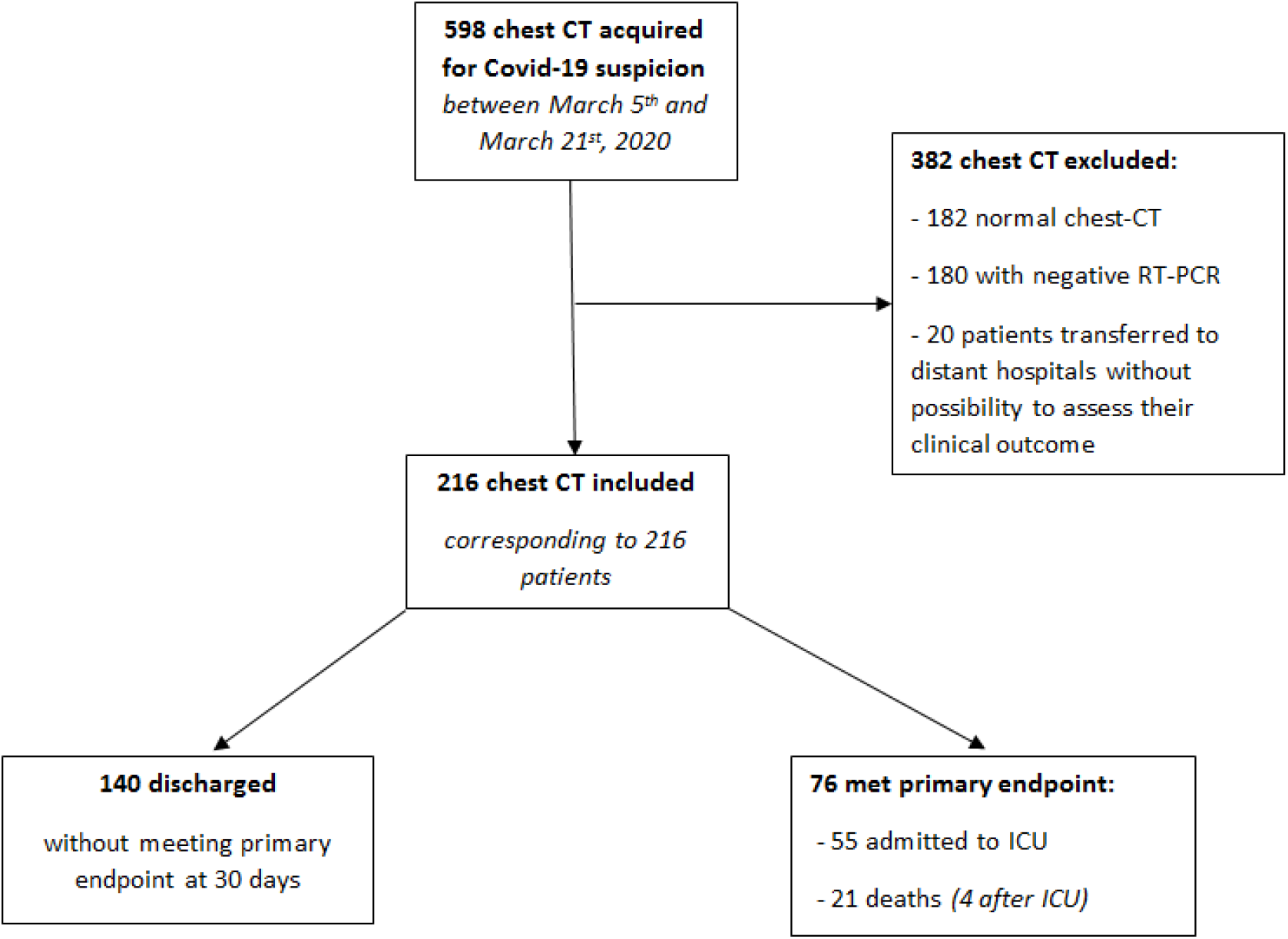
FI owchart of patients.

### Clinical Evaluation

76/216 patients (35%, 95% CI: 29.1–41.8) met the primary endpoint of death (N = 21, with 4 after an ICU stay) or mechanical ventilation (N = 55) in the 30 days following the initial chest CT.

Patients in mechanical ventilation or death group were older than patients in group without need of mechanical ventilation or death (66 years-old vs 59 years-old; p<0,001) **(Table 1)**.

**Table 1.** Patient demographic and clinical data.

|  | **All patients**<br>**(n=216)** | **Patients with**<br>**mechanical**<br>**ventilation or**<br>**death**<br>**(n=76)** | **Patients without**<br>**mechanical**<br>**ventilation or**<br>**death**<br>**(n=140)** | **P value** |
| --- | --- | --- | --- | --- |
| **Female** | 92 (43%) | 26 (34%) | 66 (47%) | p = 0.0664 |
| **Male** | 124 (57%) | 50 (66%) | 74 (53%) |  |
| **Median age in**<br>**years (SD)** | 62 (16.5) | 66 (14) | 59 (17) | p = 0.00079 |
| **Median onset**<br>**of symptoms in**<br>**days (SD)** | 6 (3.25) | 5 (3.5) | 6 (3.13) | p = 0.461 |

### CT Image Analysis

The most common subgroup of severity was 11–25% (89/216; 41%) **(Table 2)**.

**Table 2.** Patient demographic and clinical data in severity subgroups.

|  | **1-10%**<br>(n=76) | **11-25%**<br>(n=89) | **26-50%**<br>(n =37) | **51-75%**<br>(n=11) | **76-100%**<br>(n=3) |
| --- | --- | --- | --- | --- | --- |
| **Female**<br>(n=92) | 32<br>(42%) | 42<br>(47%) | 13<br>(35%) | 3<br>(27%) | 2<br>(67%) |
| **Male**<br>(n=124) | 44<br>(58%) | 47<br>(53%) | 24<br>(65%) | 8<br>(73%) | 1<br>(33%) |
| **Median age in years (SD)** | 58 (17.3) | 65 (16.2) | 62 (15.4) | 62 (14.1) | 57 (3.4) |
| **Median onset of symptoms in days (SD)** | 4 (2.4) | 7 (3.1) | 7 (3.9) | 4 (2.8) | 7 (5.9) |

Inter-reader agreement was high, with a discrepancy rate of 3% (6/216) and a weighted Cohen’s Kappa of 0.97 [94.8–99.4], endpoint was high, with a Spearman coefficient ρ of 0.87 (p = 0.05).

Occurrence of the primary outcome with regards to the lesion severity subgroups at the initial chest CT presented a major gap at a 25% threshold **(Table 3)**. A greater than 25% involvement was significantly associated with a higher risk of mechanical ventilation or death at 30 days, with a Risk Ratio of 5.00 (95% CI [3.59–6.99]).

**Table 3.**
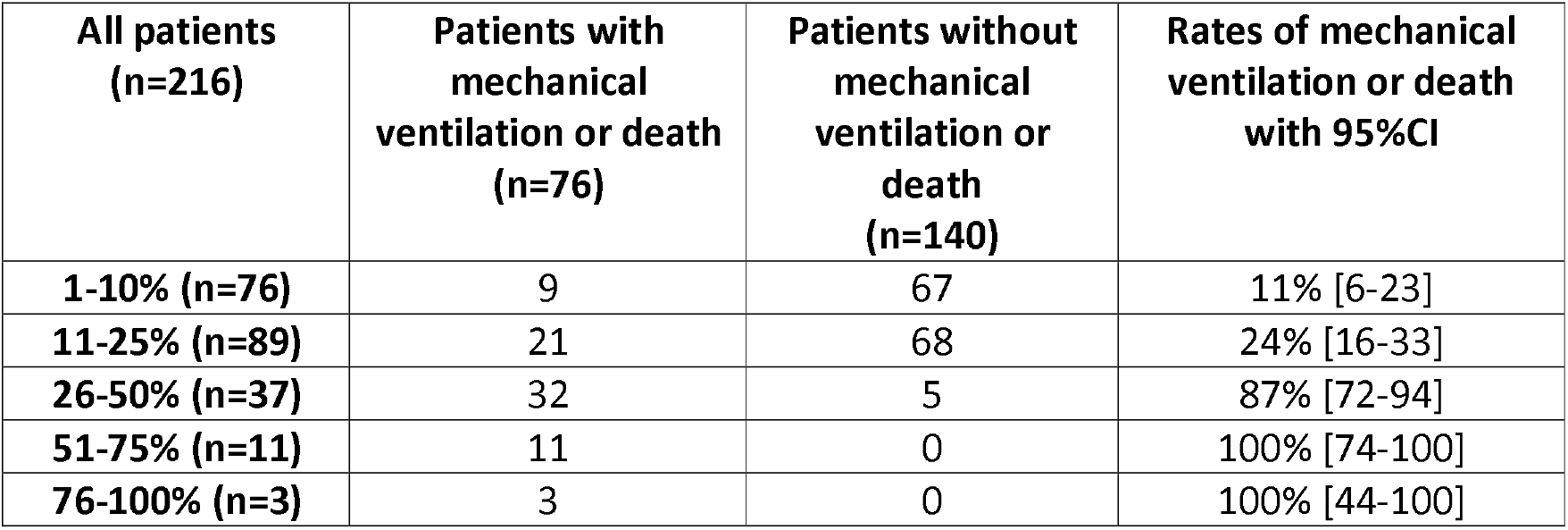
Occurrence of primary endpoint in severity subgroups.

Occurrence of ICU and/or death was 90% (95%CI [79–96]) in patients with more than 25% extensiveness versus 18% (95%CI [13–25]) in patients with less than 25% extensiveness **(Fig 2).**

**Fig 2.**
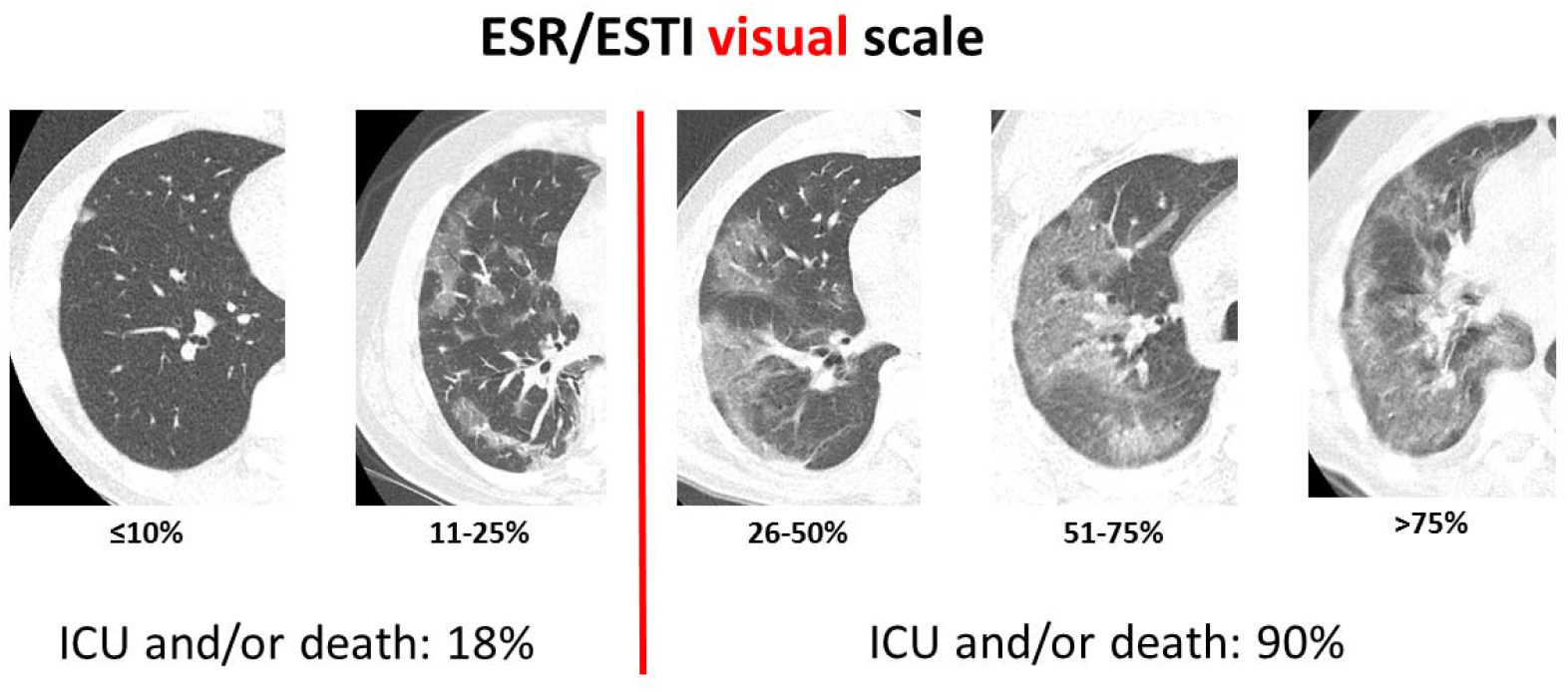
Graphical abstract.

Demonstrative examples are shown in **Figs 3 and 4**.

**Fig 3.**
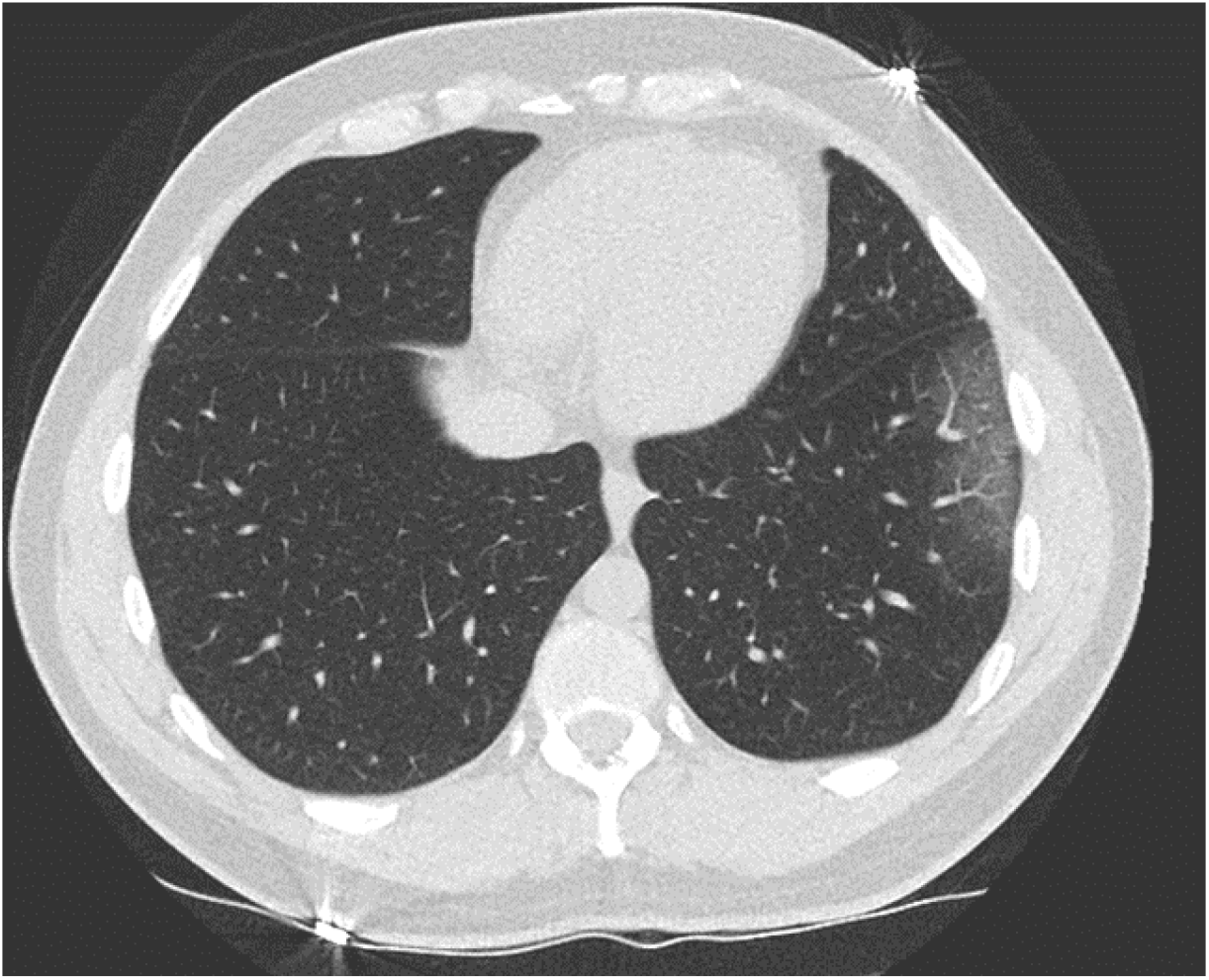
45-year-old male at day 3 of fever with acute dyspnea. Chest-CT showing limited peripheral ground-glass opacity in the left lower lobe, with a visual quantification of lesions severity of <10%. Patient was discharged home after ED admission.

**Fig 4.**
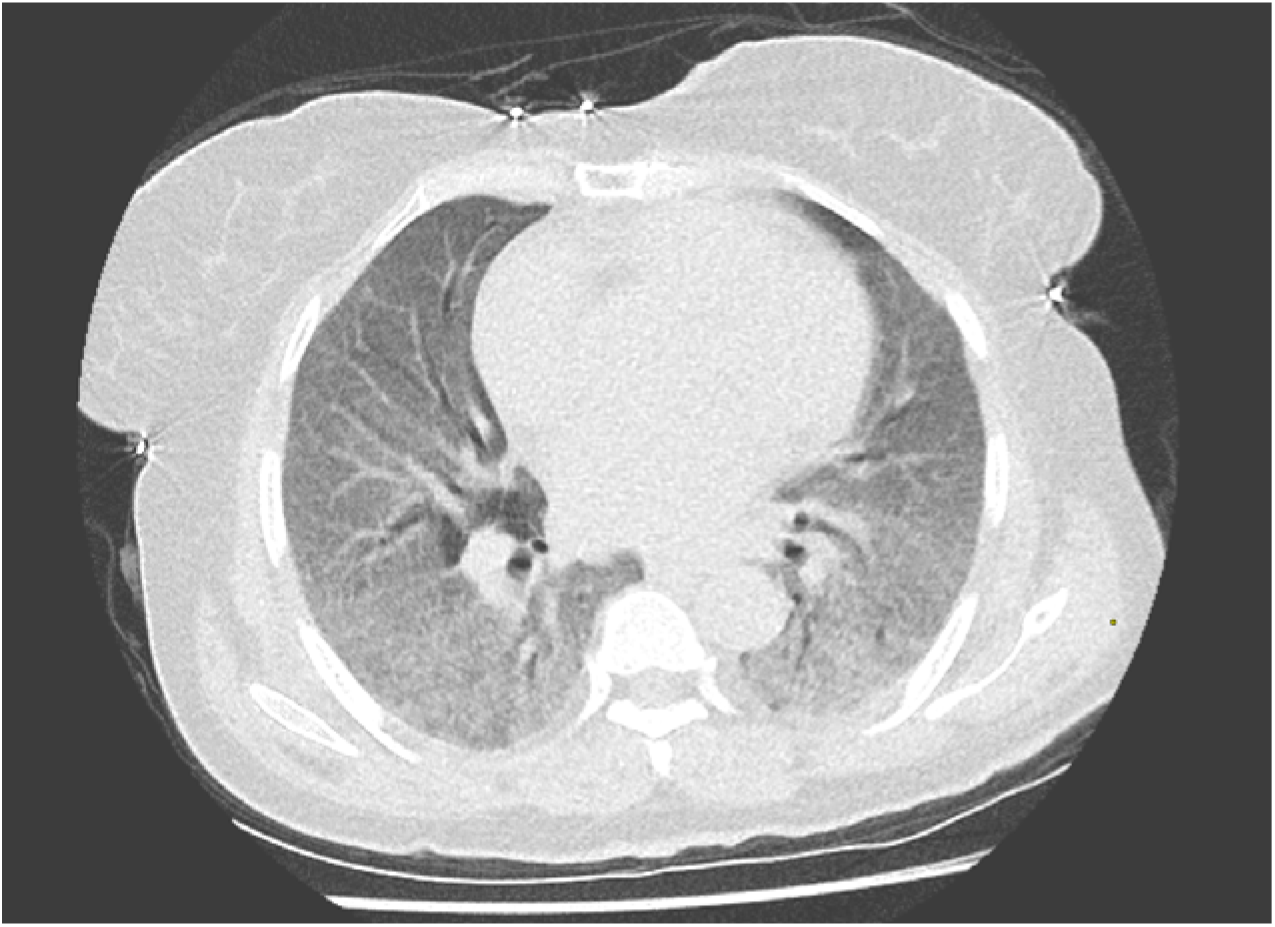
58-year-old woman at day 4 of dyspnea. Chest-CT showing extensive bilateral ground-glass opacities, with a visual quantification of lesions severity > 75%. The patient was intubated and admitted in intensive care unit few hours after CT.

## DISCUSSION

The results of the present study showed that the rate of death or mechanical ventilation in Covid-19 patients increased significantly with the degree of lesion severity at initial chest CT. Patients with more than 25% of lung parenchyma involved had a risk ratio of 5.00 for this primary endpoint. This study is, to the best of our knowledge, the first to validate the use of the ESR/ESTI CT visual assessment scale for Covid-19 lesions. Compared to semi-quantitative scores or dedicated quantification software, visual assessment has the advantage of simplicity, availability and rapidity.

Our results are in accordance with those of Colombi et al. in which a significant association between quantification of well-aerated lung parenchyma on admission chest-CT less than 73% and ICU admission or death was described (OR 5,4; p<0,001) (10), which is consistent with our proposed threshold of 25%.

Covid-19 is a novel disease, so far not fully understood and can present multiple forms and courses with a quite unpredictable outcome. Identifying prognostic factors is of major interest. In our ED workflow, chest CT is used as a triage tool for patients with moderate to severe respiratory clinical manifestations. As is, lesions severity could be used to further guide patient’s management right at the ED.

One could fear that the limiting factor of visual evaluation would be the poor reproducibility, yet we achieved a high inter-reader agreement in our work. This could be related to the fact that readers did an initial consensus training to limit this variability.

This study still presents some important limitations. First, it was a retrospective and monocentric study, with a limited number of cases. Second, other clinical data were unavailable and an independent analysis with other possible cofounding factors (12) taken into account (such as age, sex, body mass index, comorbidities, smoking status…) could not be done. Therefore, we cannot assume that CT visual scoring is an independent predictor factor of Covid-19 evolution. Third, this study was done on patients from the initial part of the epidemic wave, when pressure on ICU space was the most important and when patient’s management was not completely defined and optimized. This could partly explain the high fatality rate in our cohort. Fourth, we decided to include only patients with CT lesions, which could constitute an inclusion bias. Last, we did not perform software-based quantitative measurement to validate our visual evaluation. However, the high inter-reader agreement is in favor of the validity of this visual approach.

To conclude, this retrospective cohort confirms a correlation between visual evaluation of lesion severity at initial chest CT and the 30 days clinical outcome of Covid-19 patients, thus reinforcing the role of chest CT in ED triage. Future studies will help to refine knowledge about CT prognostic factors in Covid-19.

## Data Availability

Datas are avalaible on an Excel Word file.

## REFERENCES

1. Huang C, Wang Y, Li X, Ren L, Zhao J, Hu Y, et al. Clinical features of patients infected with 2019 novel coronavirus in Wuhan, China. The Lancet. 2020 Feb 15;395(10223):497–506.

2. Chavez S, Long B, Koyfman A, Liang SY. Coronavirus Disease (COVID-19): A primer for emergency physicians. Am J Emerg Med [Internet]. 2020 Mar 24 [cited 2020 Apr 15]; Available from: https://www.ncbi.nlm.nih.gov/pmc/articles/PMC7102516/

3. Corman VM, Landt O, Kaiser M, Molenkamp R, Meijer A, Chu DKW, et al. Detection of 2019 novel coronavirus (2019-nCoV) by real-time RT-PCR. Euro Surveill Bull Eur Sur Mal Transm Eur Commun Dis Bull. 2020;25(3).

4. Xie X, Zhong Z, Zhao W, Zheng C, Wang F, Liu J. Chest CT for Typical 2019-nCoV Pneumonia: Relationship to Negative RT-PCR Testing. Radiology. 2020 Feb 12;200343.

5. Chung M, Bernheim A, Mei X, Zhang N, Huang M, Zeng X, et al. CT Imaging Features of 2019 Novel Coronavirus (2019-nCoV). Radiology. 2020 Feb 4;295(1):202–7.

6. Yuan M, Yin W, Tao Z, Tan W, Hu Y. Association of radiologic findings with mortality of patients infected with 2019 novel coronavirus in Wuhan, China. PLOS ONE. 2020 Mar 19;15(3):e0230548.

7. Li K, Fang Y, Li W, Pan C, Qin P, Zhong Y, et al. CT image visual quantitative evaluation and clinical classification of coronavirus disease (COVID-19). Eur Radiol [Internet]. 2020 Mar 25 [cited 2020 Apr 15]; Available from: https://doi.org/10.1007/s00330-020-06817-6

8. Zhao W, Zhong Z, Xie X, Yu Q, Liu J. Relation Between Chest CT Findings and Clinical Conditions of Coronavirus Disease (COVID-19) Pneumonia: A Multicenter Study. AJR Am J Roentgenol. 2020 Mar 3;1–6.

9. Yang R, Li X, Liu H, Zhen Y, Zhang X, Xiong Q, et al. Chest CT Severity Score: An Imaging Tool for Assessing Severe COVID-19. Radiol Cardiothorac Imaging. 2020 Mar 30;2(2):e200047.

10. Colombi D, Bodini FC, Petrini M, Maffi G, Morelli N, Milanese G, et al. Well-aerated Lung on Admitting Chest CT to Predict Adverse Outcome in COVID-19 Pneumonia. Radiology.2020 Apr 17;201433.

11. Revel M-P, Parkar AP, Prosch H, Silva M, Sverzellati N, Gleeson F, et al. COVID-19 patients and the radiology department – advice from the European Society of Radiology (ESR) and the European Society of Thoracic Imaging (ESTI). Eur Radiol [Internet]. 2020 Apr 20 [cited 2020 May 8]; Available from: https://doi.org/10.1007/s00330-020-06865-y

12. Richardson S, Hirsch JS, Narasimhan M, Crawford JM, McGinn T, Davidson KW, et al. Presenting Characteristics, Comorbidities, and Outcomes Among 5700 Patients Hospitalized With COVID-19 in the New York City Area. JAMA [Internet]. 2020 Apr 22 [cited 2020 May 14]; Available from: https://jamanetwork.com/journals/jama/fullarticle/2765184

